# “We worked as a team”: Frontline providers’ experiences of a multi-cadre training initiative for early identification, care, and referral for children with developmental disabilities in Kenya

**DOI:** 10.64898/2026.08.18.26360684

**Authors:** Beatrice Mkubwa, Amina Abubakar, Melissa Washington-Nortey, Carophine Nasambu, Samlee Nyambu, Eva Mwangome, Walter Kisangi, Eddie Chengo, Nancy Githinji, Tsegereda Haile Kifle, Mekdes Demissie, Fikirte Girma, Marit Sijbrandij, Charles R. Newton, Rosa A. Hoekstra, Vibian Angwenyi

**Author notes:** Corresponding author: (BM).

## Abstract

Children with developmental disabilities remain among the most underserved globally, with significant delays in early identification and access to care. In Kenya, limited capacity in the frontline workforce further disrupts the timely recognition and management of DDs. This study evaluated the impact of a multi-cadre capacity-strengthening training intervention on the knowledge and practice of healthcare workers (HCWs) and community support workers (CSWs) (including community health promoters, teachers, and caregivers of children with disabilities) to improve early identification, assessment, care, and referral of children with DDs in Kenya.

We conducted a sequential mixed-methods study and collected data between 2023 and 2025. Quantitative measures included sociodemographic surveys for all participants (CSWs and HCWs) and pre- and post-knowledge assessments using a 15-item Mental Health Gap Action Programme Intervention Guide (mhGAP-IG)-based questionnaire administered only to HCWs participating in the training. Qualitative data were obtained through focus group discussions with CSWs (n=5, 58 participants) and HCWs (n=5, 48 participants) to explore training experiences, perceived skill gains, and implementation experiences. Quantitative analysis used descriptive statistics and Wilcoxon signed-rank tests, while qualitative data were analysed thematically.

A total of 321 frontline providers were trained. Ninety-seven HCWs from 25 public health facilities received DDs training based on the WHO mhGAP-IG module, while 224 CSWs received a DD-focused co-designed training on community-based early identification and referral. Among HCWs with pre-post assessments (n=70), knowledge scores improved significantly (mean change: +0.86, p < 0.001), and the proportion scoring ≥12 increased from 51% to 75%. Qualitative findings post-implementation indicated strengthened capacity in developmental milestone assessment, identification of DDs, and improved referral across the community, facility and other related sectors. Both cadres reported improved confidence in addressing myths and misconceptions related to DDs, coordinated teamwork, and improved caregiver engagement during assessment, and referrals driven by improved provider confidence and more supportive communication following the training. Increased workload, limited time for assessments, and limited resources were reported as challenges.

This evaluation of a multi-cadre training model demonstrated improved knowledge, skills, and confidence among CSWs and HCWs, improving early identification, care, and referral practices for children with DDs. Sustaining these gains will require ongoing supervision, integration into routine workflows, strengthened referral systems, and continued investment in frontline workforce development within primary care settings.

## Introduction

Developmental disorders (DDs) are conditions that arise during the developmental period and affect the acquisition of cognitive, motor, language, communication, or social skills, leading to functional impairment. Children represent one of the most underserved populations in health systems globally [1]. Hundreds of millions of children live with DDs worldwide, with the greatest burden borne by low- and middle-income countries (LMICs), where access to early identification, intervention, and care remains limited [1–4]. Delays in diagnosis and intervention are associated with long-term adverse outcomes across health, education, and social domains, underscoring the importance of early detection and timely care during early childhood [5], which represents a critical window during which intervention can substantially improve developmental outcomes [6].

In Kenya, as in many sub-Saharan African countries, systemic constraints hinder timely identification and care for children with DDs. Shortages of specialist providers, limited training in developmental screening among frontline workers, weak referral systems, and fragmented service delivery contribute to prolonged delays in accessing appropriate services for early developmental concerns [7–11]. Stigma, low caregiver awareness, and harmful cultural beliefs (e.g., attributing DDs to witchcraft, curses, ancestral spirits, or maternal wrongdoing) worsen these challenges, sometimes leading families to hide children with DDs or disengage from care [12–14]. Emerging evidence from both urban and rural Kenya highlights persistent unmet need, inconsistent follow-up, and inequities in access to neurodevelopmental services, particularly among socioeconomically disadvantaged populations [15, 16]. These gaps represent a missed opportunity to intervene during a period of heightened developmental plasticity.

Frontline workers across health, education, and the community play a central role in early identification, care, referral, and family support within Kenya’s Primary Healthcare (PHC)– oriented Primary Care Networks [17]. Community Health Promoters (CHPs) identify developmental concerns at the household level, teachers detect early learning or behavioural difficulties in school settings, and nurses and clinical officers provide the first formal clinical point of contact at PHC facilities [18]. Caregivers serve as day-to-day monitors of children’s development and drive help-seeking, reinforcing continuity and coordination of care across the health, education and community sectors [8]. However, many frontline workers report insufficient knowledge, limited confidence, and inadequate skills to identify or manage DDs, resulting in missed early warning signs and weak linkages between households, schools, and health services [19].

Task-sharing approaches, where non-specialist providers deliver elements of care traditionally performed by specialists, have been promoted as effective strategies to bridge the specialist shortage in LMICs [20–22]. Evidence from India, Ethiopia, and other LMICs indicates that structured training, job aids, and supportive supervision can improve developmental screening and management of DDs [6, 20, 23]. Many capacity-strengthening initiatives concentrate on facility-based healthcare workers (HCWs), often excluding teachers, CHPs, and caregivers of children with disabilities, despite their crucial roles in the early detection and care pathway for DDs [24–26]. As a result, challenges persist in multisectoral coordination of early identification at the community and school levels, linkage to care, and meaningful caregiver engagement, defined here as caregivers’ active, informed participation in identification, care decisions, and management [27, 28].

Global frameworks emphasise the need for integrated and multisectoral systems to support early childhood development. The Mental Health Gap Action Programme Intervention Guide (mhGAP-IG) promotes developmental assessment within PHC. At the same time, the Nurturing Care Framework highlights the shared responsibilities across caregivers, communities, education, and health sectors [29, 30]. In addition, the Chronic Care Model emphasises the importance of coordinated, person-centred care for long-term conditions such as DDs, further reinforcing the need for sustained follow-up and strong linkages across community and health systems [31–33]. Evidence from Uganda’s Baby Ubuntu trial [34], a child psychosocial stimulation programme in Bangladesh [35], and a review of school-based intervention for children with DDs in LMICs [36], shows that mental health-related community initiatives integrating caregivers, educational settings, and PHC training are more effective than single-sector approaches. Nevertheless, in Kenya and similar African contexts, empirical evidence remains limited on how strengthening frontline worker capacity across community, educational, and PHC settings can improve early identification, care, and referral for children with DDs [8, 37].

To address this gap, the present study evaluates the impact of an integrated, multi-cadre capacity-strengthening training intervention on knowledge and practice to improve early identification, care, and referral pathways for children with DDs in Kenya.

## Methods

### Study design

This study employed a sequential mixed-methods design to evaluate a two-tier capacity-strengthening training intervention for frontline providers involved in early identification, care, and referral of children with DDs in Kenya [38]. A sociodemographic survey was administered to all participants; quantitative data were collected to assess changes in HCWs’ knowledge following the training (administered only to HCWs), followed by focus group discussions (FGDs) to explore participants’ experiences of the training and post-training implementation. Data were collected between 2023 and 2025.

The study was embedded within the SPARK project (SuPporting African communities to increase Resilience and mental health of Kids with developmental disabilities) [13], an international research collaboration implemented in Kenya and Ethiopia to improve the outcomes of children with DDs and their caregivers.

### Study setting

The study was conducted in Kilifi County, a predominantly rural coastal region, and in informal settlements in Nairobi County, Kenya’s capital city. Training and data collection were conducted across 25 public health facilities that served as catchment areas for the SPARK project shown in figure 1 below and the sub-counties they fall under. These sites provided a mix of dispensaries, health centres, and hospitals, linked to surrounding communities and schools through existing PHC and community health structures.

**Figure 1.**
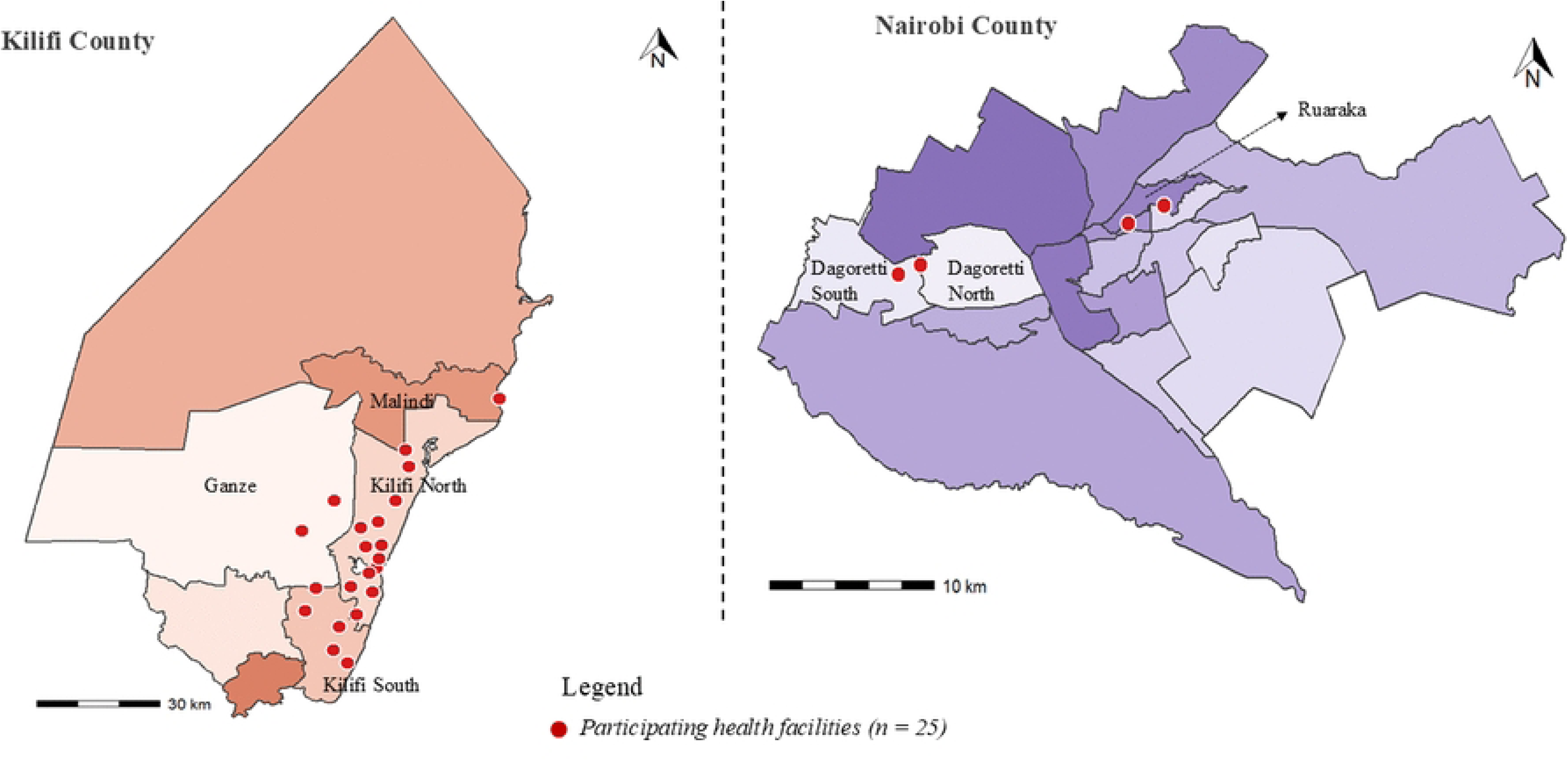
Map of the 25 public health facilities and their catchment areas included in the SPARK project, across Nairobi and Kilifi Counties, Kenya.

### Participants and sampling

Participants comprised frontline providers operating at community and PHC levels who took part in the SPARK training intervention. Two cadres were included: a) ‘Community support workers’ (CSWs), an umbrella term, endorsed following stakeholder consultations, for trusted community-based resource personnel involved in identifying and referring children with developmental concerns. This includes Ministry of Health-supported CHPs, teachers, and caregivers of children with DDs who had lived experience and were engaged in community support roles; and b) HCWs, facility-based PHC providers involved in the assessment and care of children with DDs, including nurses, clinical officers, and other frontline staff.

Individuals who could take on the CSWs’ role were identified before the training based on whether they had regular contact with children and caregivers in households, schools, or the community. Eligibility for CSWs was therefore determined by professional responsibilities or lived experience that positioned them to support early identification of DDs. HCWs were eligible if they were involved in routine PHC service delivery and selected by county health authorities and facility managers to participate in the WHO mhGAP-IG training on DDs. For the qualitative component of the study, participants were invited purposively. Specifically, individuals who completed the relevant training and remained actively engaged during the study implementation, prioritising those who represented a wide range of demographic backgrounds, were invited for the FGDs. No additional exclusion criteria were applied.

### Training intervention overview

The intervention adopted a two-tier, task-sharing model with separate but complementary training packages for CSWs and HCWs, reflecting their distinct roles in the early identification and referral pathway.

### Training for community support workers

CSWs participated in a three-day training designed to build foundational knowledge and practical skills related to child development and DDs. The training content included: basic child developmental domains and milestones; early warning signs of developmental delays and disabilities; differentiating developmental delay from physical disabilities; using non-stigmatising language and respectful communication with caregivers; and an overview of referral pathways and available services.

The training used a participatory approach, combining short lectures, locally adapted case studies, images, milestone charts, group discussions, and role plays. Content was refined iteratively based on participant feedback throughout the training. CSWs were expected to apply these skills during household visits, school interactions, and community engagement to identify children with possible developmental concerns and facilitate timely linkage to health facilities for assessment.

### Training for healthcare workers

HCWs completed a three-day, competence-focused training based on the WHO mhGAP-IG principles, with specific emphasis on DDs. The training aimed to strengthen HCWs’ capacity to assess and classify DDs, provide basic psychosocial interventions, including psychoeducation and counselling, and make appropriate/timely referrals for specialised assessment and care. Training content covered early identification, structured developmental assessment, guidance and counselling, and standardised referral procedures. Materials were tailored to the Kenyan PHC context and incorporated DSM-5-informed principles relevant to DDs. Training methods included lectures, case-based discussions, and interactive exercises designed to strengthen practical assessment skills. HCWs were supported to use structured tools and job aids to guide evaluation and communication with caregivers.

### Supervision, Mentorship, and Follow-Up

Following training, regular review meetings, supervision visits, and continuous feedback were implemented across both cadres. These activities supported mentorship, ongoing problem-solving, and coordination between community-and facility-based providers. The intention was to strengthen referral pathways and communication across households, schools, communities, and health facilities.

### Data collection procedures

#### Quantitative data collection

A structured, self-administered questionnaire was used to capture participants’ sociodemographic characteristics, including age, sex, cadre, years of experience, and facility type. Knowledge related to child developmental, mental, and behavioural disorders was assessed using a 15-item multiple-choice questionnaire adapted from the WHO mhGAP-IG child and adolescent mental and behavioural disorders module. Each item had up to four response options, with only one correct response. Correct responses were scored as one point, yielding a total score range of 0-15, with higher scores indicating greater knowledge. The knowledge questionnaire was administered before (pre-test) and immediately after (post-test) completion of the HCW training.

#### Qualitative data collection

Qualitative data were collected approximately 3 months post-training through 10 FGDs, five with CSWs and five with HCWs. A semi-structured FGD guide explored participants’ perceptions of the training, perceived knowledge and skill gains, experiences in routine practice, and the contextual factors influencing early identification and referral of children with DDs. FGDs were conducted in English or Kiswahili by experienced qualitative researchers, lasted approximately 60-90 minutes, and were audio recorded with participants’ consent. Field notes were taken during discussions. Audio recordings were transcribed verbatim and translated into English where necessary.

### Data management and analysis

#### Quantitative analysis

Data were cleaned and analysed using R (version 4.4.1). Sociodemographic characteristics were summarised using descriptive statistics. Knowledge scores were summarised as means, standard deviations, medians, and interquartile ranges. Pre-post changes in knowledge were calculated as the difference between post-test and pre-test scores. Because the data were not normally distributed, the Wilcoxon signed-rank test was used to evaluate differences in knowledge scores. Statistical significance was set at p < 0.05. Exploratory stratified analyses were conducted by site, age group, gender, cadre, facility type, and prior training exposure.

#### Qualitative analysis

Qualitative data were analysed using thematic analysis following Braun and Clarke’s six-step approach [39]. Two researchers independently reviewed transcripts and developed an initial coding framework, combining inductive codes derived from the data with deductive codes informed by the study objectives and FGD guides. Coding discrepancies were discussed and resolved through consensus, with input from a third senior qualitative researcher when needed. All transcripts were coded using NVivo-Lumivero^©^ version 12. Themes were reviewed, refined, and compared across cadres and study sites. Representative participant quotes were selected to illustrate key findings.

Qualitative and quantitative findings were integrated during interpretation using a triangulation, with comparisons across datasets identifying areas of convergence, complementarity, dissonance, and silence, thereby facilitating the development of overarching meta-themes and the iterative exploration of key findings across methods.

### Ethical considerations

Ethical approval for the study was obtained from The Aga Khan University Institutional Scientific and Ethics Review Committee (ISERC) Ref: 2021/ISERC-99 (V4, 26-Feb-2024), Kenya Medical Research Institute Scientific and Ethics Review Unit (SERU) Ref: KEMRI/SERU/CGMR-C/272/4872 (V1.3, 26-Jan-24), Kings College London Ethics Approval Ref: RESCM-24/25-18398, and National Commission for Science, Technology and Innovation (NACOSTI), Kenya Ref: NACOSTI/P/24/33550. Research permits were also obtained from the health authorities of Nairobi and Kilifi Counties. Informed consent was sought from all participants prior to data collection. Although institutional permission was granted to access facilities and frontline providers, participation in the training and all study activities was voluntary. All data were de-identified and securely stored, with access restricted to the research team.

## Results

A total of 321 participants (224 CSWs and 97 HCWs) completed the training and were included in the overall analysis. Sociodemographic data were available for all trained CSWs and HCWs (Tables 1 and 2). Knowledge assessment data were collected from HCWs who had been trained only on the WHO mhGAP module. A subsample of 106 trained frontline providers participated in the qualitative evaluation component of the study, including 58 (25.9%) CSWs and 48 (56.5%) HCWs.

**Table 1.** Sociodemographic and professional characteristics of community support workers (n = 224)

| *Category* | *n (%) or Mean (SD)* |
| --- | --- |
| *Site* |  |
| Nairobi | 78 (34.8) |
| Kilifi | 146 (65.2) |
| *Gender* |  |
| Male | 58 (25.9) |
| Female | 166 (74.1) |
| *Age (years)* | M=45.2 (SD 10.2) |
| *Education level* |  |
| Primary | 87 (38.8) |
| Secondary | 96 (42.9) |
| Tertiary or above | 41 (18.3) |
| *Main occupation\** |  |
| Community health promoter | 180 (80.4) |
| Teacher (regular or special school) | 15 (6.7) |
| Unemployed | 12 (5.4) |
| Other | 12 (5.4) |
| Missing | 5 (2.2) |
| *Other positions held\** |  |
| Local leader | 124 (55.4) |
| Community health promoter | 43 (19.2) |
| Caregiver of child with DD/Lived experience | 19 (8.5) |
| Teacher | 12 (5.4) |
| Other | 21 (9.4) |
| Missing | 5 (2.2) |
| *Type of organisation* |  |
| Dispensary | 132 (58.9) |
| Health centre | 48 (21.4) |
| School | 16 (7.1) |
| Home / unemployed | 21 (9.3) |
| Other | 7 (3.1) |
| *Years of work experience* |  |
| Mean (SD) | 9.6 (7.3) |
| *Prior experience with children/families with disabilities* |  |
| Yes | 127 (56.7) |
| No | 97 (43.3) |
| *Previous general disability training* |  |
| Yes | 86 (38.4) |
| No | 136 (60.7) |
| Missing | 2 (0.9) |
| *Previous DDs training* |  |
| Yes | 57 (25.4) |
| No | 164 (73.2) |
| Missing | 3 (1.3) |

*Note: Percentages may not total 100 due to rounding. Missing data across variables ranged from 0–2.5%.*
*\*Main occupation and other positions held may overlap, as some participants held more than one role.*

**Table 2.** Sociodemographic and professional characteristics of trained healthcare workers (n = 85)

| *Category* | *n (%) or*<br>*Mean (SD)* |
| --- | --- |
| *Site* |  |
| Nairobi | 43 (44.3) |
| Kilifi | 54 (55.7) |
| *Gender* |  |
| Female | 78 (80.4) |
| Male | 19 (19.6) |
| *Age (years)* | M=38.4 (SD 7.8) |
| *Occupation\** |  |
| Nurse | 56 (57.7) |
| Clinical Officer | 18 (18.6) |
| Allied mental health professional | 10 (10.3) |
| Allied health / administrative | 9 (9.3) |
| Other | 4 (4.1) |
| *Facility type* |  |
| Hospital | 45 (46.4) |
| Dispensary | 32 (33.0) |
| Health Centre | 18 (18.6) |
| EARC* | 2 (2.1) |
| *Position held\** |  |
| Leadership/management | 39 (40.2) |
| Clinical services providers | 37 (38.1) |
| Administrative/support | 15 (15.5) |
| Programme/coordination | 6 (6.2) |
| *Previous disability-related training* |  |
| No | 80 (82.5) |
| Yes | 17 (17.5) |
| *Previous DDs training* |  |
| No | 79 (92.9) |
| Yes | 6 (7.1) |
| *Years of work experience* | M=10.3 (SD 7.9) |

\*Occupation and positions held may overlap, as some participants held more than one role.
\*Educational Assessment and Resource Centre

### Sociodemographic characteristics of the trained CSWs

Of the 224 CSWs trained (Table 1), the majority (65.2%) were from Kilifi, and one-third were from Nairobi. The median age of the CSWs was 45.5 years (IQR = 38-52), and nearly half were aged 35-49 years. Most participants were female (74.1%) and had attained at least secondary-level education (42.9%). Professionally, most participants were CHPs (80.4%) working primarily in public health facilities (80.3%), with an average of 9 years of work experience (IQR = 5-12).

#### Pre-training exposure to disability and developmental disabilities among CSWs

Just over half of the CSWs (56.7%) reported previous experience working with children or families affected by disabilities. Site differences were observed, with a higher proportion reporting prior experience in Nairobi (83.3%) compared to Kilifi (42.5%). Approximately 38.4% (n=86) had received some form of general disability-related training. Among those trained, 36 had been trained in recognising symptoms, 20 in applying behavioural activation directly with the child, and 28 in supporting caregivers of children with general disabilities; training information was missing for four participants. The median time since participants last received general disability training was 2 years (IQR = 0.8-30).

Only 38.4% (n=86) of participants had received some form of general disability-related training. Among those trained, 36 had been trained to recognise symptoms, 20 to apply behavioural interventions directly with the child, and 28 to support caregivers. Previous general disability training was more common in Nairobi (65.4%) than in Kilifi (24.0%). A smaller proportion (25.4%, n=57) reported receiving prior training specifically focused on DDs. Among these, 25 had received training on recognising DD symptoms, 19 on applying behavioural interventions directly with the child, and 12 on supporting caregivers of children with DDs; one response was missing. The median time since the last DD-specific training was 1 year (IQR = 0.2–3.0).

For DDs specifically, 25.4% (n=57) of participants reported prior training. Among those with training, 25 had been trained to recognise symptoms, 19 to apply behavioural interventions directly with the child, and 12 to support caregivers. As with other indicators, prior developmental disability training was higher in Nairobi (47.4%) compared to Kilifi (13.7%).

## Sociodemographic characteristics of HCWs

Ninety-seven HCWs received the mhGAP training (Table 2), with 55.7% drawn from Kilifi and the rest from Nairobi. Most trainees were female (82.4%), with an average age of 38.4 years. Almost 50% of trained HCWs were stationed in hospitals, 36.7% in dispensaries, and 12.9% in health centres. Their professional roles included nurses (55.3%), clinical officers (17.6%), and other occupations (e.g., administrative and allied health staff) (27.1%). When collapsed into broader categories, 24.7% held leadership positions as heads or managers. Experience levels varied, with approximately half of the participants reporting less than 10 years of professional experience (50%).

### Pre-training exposure to disability and developmental disabilities among HCWs

Most healthcare workers had not received any disability-related training before the intervention. A total of 17.5% (n = 17) reported previous general disability training, while 82.5% (n = 80) had not. Among those with prior training, 11 HCWs (11.3%) had been trained in general disability assessment, 4 (4.1%) in providing direct support to persons or children with disabilities, and 2 (2.1%) in supporting caregivers. One respondent did not specify the type of training received. The median time since the last general disability training was 3.5 years (IQR: 0–10.8 years).

Training specifically focused on DDs was less common. Only 9.3% (n = 9) of HCWs reported having received DD-specific training, compared to 90.7% (n = 88) with no such prior exposure. Of those trained, 8 HCWs (8.2%) had received training in recognising or assessing DD symptoms, while 1 (1.0%) had been trained to provide direct support to a child with DDs. The median time since the last DD-specific training was 2 years (IQR: 0.8–10 years).

### Knowledge scores pre- and post-training

At baseline (pre-test), participants scored a mean of 11.5 (SD 1.83), with a median of 12 (IQR: 10–13). At post-test, the mean knowledge score increased to 12.4 (SD 1.34), with a median of 12.5 (IQR 11.8–13) (Table 3). The proportion of participants scoring ≥12 increased from 51.2% at pre-test to 75% at post-test, indicating improved overall knowledge. Ceiling effects were minimal, with only 3.6% and 4.2% achieving the maximum score of 15 on the pre- and post-tests, respectively.

**Table 3.**
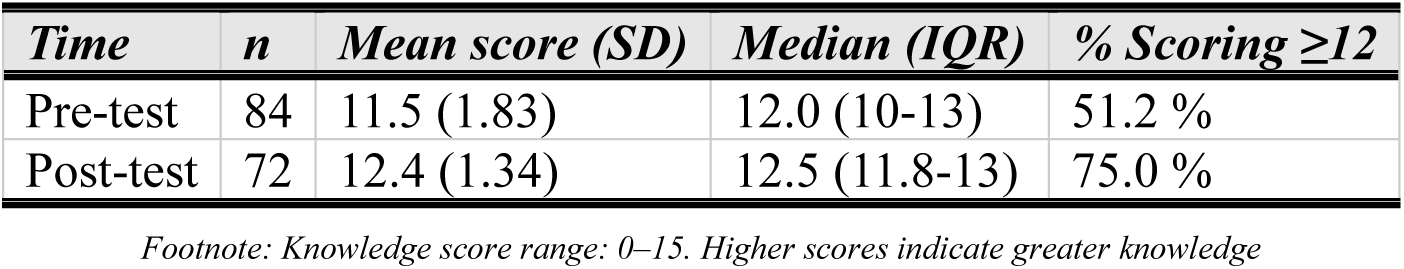
Knowledge scores among trained healthcare workers before and after training.

| *Time* | *n* | *Mean score (SD)* | *Median (IQR)* | *% Scoring $\geq 12$* |
| --- | --- | --- | --- | --- |
| Pre-test | 84 | 11.5 (1.83) | 12.0 (10-13) | 51.2 % |
| Post-test | 72 | 12.4 (1.34) | 12.5 (11.8-13) | 75.0 % |

*Footnote: Knowledge score range: 0–15. Higher scores indicate greater knowledge*

### Pre-post change in knowledge

Among the 70 participants with complete paired data, the mean change in knowledge score was 0.86 points (SD 1.89), with a median increase of one point (IQR –0.75 to 2.75). Over half of participants (54.3%) showed improvement, while 20% demonstrated no change, and 25.7% experienced slight declines. A Wilcoxon signed-rank test confirmed that the increase in knowledge was statistically significant (V = 1204.5, p < 0.001). Baseline pre-test scores did not differ significantly between participants who completed the post-test assessment and those lost to follow-up, suggesting limited evidence of selective attrition (Wilcoxon rank-sum test: W = 6.5, p = 0.165).

### Stratified knowledge gains

Exploratory analyses indicated similar improvements across sites, with mean pre-post changes of 0.88 in Nairobi and 0.84 in Kilifi. Knowledge gains were observed across all age groups, with the largest mean increase among participants aged 50+ (1.2 points). By professional role, clinical officers/doctors gained an average of 1.17 points, nurses 1.06 points, and heads/managers 0.6 points. Female participants demonstrated slightly higher mean gains (1.01) than male participants (0.13). Gains were comparable across facility types and prior training exposure, although participants with previous training generally started from higher baseline scores.

### Qualitative findings

#### Characteristics of qualitative FGD participants

Among CSWs, the mean age was 44 years (SD=11; median=45), and the mean work experience was 11.5 years (SD=7.9; median=10.5). Most CSWs were female (79%) and primarily worked as CHPs (81%) linked with a government or public health facility. The majority were based in Nairobi County (67%).

HCWs participating in the FGDs were slightly younger, with a mean age of 37 years (SD=8; median=36) and an average of 11 years of professional experience (SD=7; median=9). Females comprised 75% of the HCWs in the qualitative sample. Nurses represented the largest cadre (58%), followed by clinical officers (15%), with smaller numbers of allied health and administrative professionals. Detailed sociodemographic characteristics of qualitative FGD participants are provided in Supplementary File 1 (CSWs) and 2 7B (HCWs).

### Perceived impact of training on DDs

Thematic analysis identified five major themes across CSWs’ and HCWs’ group discussions reflecting participants’ experiences of the training and its application in practice. A summary of findings are provided in Box 1. Findings were largely consistent across cadres and study sites, although the scope of practice varied by role. Each theme is explained in detail below, including illustrative quotes. The quote source information indicates whether the data were from FGDs with HCWs or CSWs, in Nairobi (NBO) or Kilifi (KLF), and the participant number assigned at the beginning of the FGDs (e.g., P1, P2, P3). Detailed quotes are also provided in Supplementary File 3.

#### Box 1. Summary of Key Qualitative Findings

##### Healthcare Workers

*a)* Reported improved ability to identify and differentiate developmental disabilities (ASD, ADHD, GDD, cerebral palsy)
*b)* Increased confidence in conducting assessments and making timely referrals to other health facilities
*c)* Strengthened use of structured tools and job aids to guide diagnosis
*d)* Reduced stigma and more empathetic engagement with children and caregivers
*e)* Greater collaboration within facilities through peer support, CMEs, and mentorship
*f)* Knowledge cascade extended to other departments and other non-trained HCWs
*g)* Identified gaps in managing paediatric mental health, behavioural problems, and linking families to social support
*h)* Recommended longer, more practical, and inclusive training sessions with regular follow-up

##### Community Support Workers

*a)* Gained new knowledge on developmental milestones and early warning signs
*b)* Improved recognition of developmental delays versus disabilities
*c)* Enhanced confidence in engaging caregivers and providing guidance
*d)* Contributed to early identification of children with developmental concerns in the community and timely linkage to health facilities for formal assessment
*e)* Shared knowledge within families and neighbourhoods, creating a ripple effect
*f)* Reduced stigma and dispelled myths about developmental disabilities
*g)* Increased awareness of referral pathways and local support services through a study co-developed resource kit/information guide
*h)* Recommended expanded coverage, practical tools, and stronger community engagement strategies

### Training on DDs: approach, experiences, and recommendations

#### Participatory and interactive training approach

Participants across both cadres valued the interactive, case-based training approach, which facilitated active engagement and the practical application of knowledge.

> *The sessions were very engaging because we were not just listening. The case discussions made it easier to relate the content to what we see in the clinic daily*. (P1_HCW_FGD04_NBO)

CSWs similarly valued the opportunity to learn collaboratively:

> *Everyone participated, and sharing our experiences helped us understand better*. (P7_CSW_FGD05_KLF)

#### Supportive facilitation

Training facilitators were described as approachable and patient, creating a learning environment that encouraged participation and enabled effective learning.

> *The facilitators encouraged questions, which made it easier to clarify confusing areas, especially regarding assessment and classification*. (P6_HCW_FGD01_KLF)
>
> *The facilitators were patient and did not rush us; they made sure everyone understood*. (P10_CSW_FGD03_NBO)

### Relevance and depth of training content

Participants highlighted that the training was comprehensive and addressed gaps in prior knowledge on DDs in greater depth than in previous learning experiences.

> *Although the training was intensive, it covered many areas that we had never been trained on practically, especially DDs*. (P10_HCW_FGD02_KLF)

### Recommendations for strengthening training

Both cadres identified areas for improvement, particularly the need for additional training on complex and co-occurring conditions with DDs. A HCW said:

> *Some conditions are still difficult to assess, especially intellectual disability and learning problems, and we need more guidance on that*. (P9_HCW_FGD02_KLF)
>
> *We touched briefly on autism, but there are still complex conditions like …epilepsy that we need to understand better*. (P12_CSW_FGD05_KLF)

Participants recommended extending the training duration and expanding the range of DD topics covered to allow deeper learning and greater practical application.

### Improved knowledge and competencies in the identification and management of DDs

#### First time experiences with DDs

Across both sites (Kilifi and Nairobi), many participants from both cadres described the training as their first formal training on DDs. They stated that it changed their perception of children with developmental challenges within their communities.

> *The training was an eye opener. I did not know there were so many children with developmental challenges hidden in the community*. (P1_CSW_FGD02_KLF)

#### Improved understanding of developmental milestones and trajectories

Both CSWs and HCWs reported increased capacity to assess a child’s behaviour with age-appropriate expectations and refer, enhancing their ability to identify early warning signs of DDs.

> *We learnt about child growth and development. When you see two children of the same age, and one is doing things differently, you can tell there might be a developmental problem*. (P1_CSW_FGD05_KLF)

HCWs similarly described improved capacity to monitor development.

> *The training helped us better understand the developmental stages. Before, we were not very keen on age-specific milestones, but now it is easier to know when something is not progressing as expected*. (P6_HCW_FGD05_KLF)

#### Recognition, assessment, and management of DDs

Before the training, many participants reported having associated DDs with visible physical impairments. They described the training to have broadened their understanding to include disabilities that are less visible physically, such as speech delays, learning difficulties, and atypical social behaviour, as well as co-occurring neurodevelopmental conditions.

> *Some children look physically normal, but they have challenges with learning or socialising, and now we can recognise that*. (P6_CSW_FGD05_NBO)
>
> *We learnt that some neurological conditions could present together with developmental disabilities, and this helped us not to miss important signs when assessing children*. (P12_HCW_FGD04_NBO)

HCWs reported increased confidence in conducting structured assessments, which became easier to apply with practice. Improved documentation and diagnostic clarity were also noted. HCWs described increasing their use of assessment tools, for example, the DSM-5-informed assessment tool, over time, enabling more structured assessments and clearer communication with caregivers.

> *At first, using the tools felt difficult and time-consuming, but with practice, it became easier. Now I am confident when doing assessments because I know what to look for and how to explain the findings to the caregiver*. (P2_HCW_FGD01_NBO)
>
> We also started documenting these cases better, which helped us realise that developmental disabilities were more common than we previously thought. (P3_HCW_FGD02_KLF)

CSWs reported greater confidence during household visits and improved differentiation between delay and disability.

> *Before the training, many of these children were hidden in homes, but now we can identify them during visits*. (P2_CSW_FGD04_NBO)

Beyond assessment, some HCWs reported the capacity to provide basic management, including psychoeducation, guidance and counselling, and regular monitoring.

> *We can now provide caregivers with guidance on stimulation and simple interventions while they wait for referrals, rather than just sending them away*. (P9_HCW_FGD05_KLF)

HCWs also described making informed decisions regarding referral pathways:

> *Even when I am not completely sure, the training helped me know the next step, whether to start basic counselling, monitor the child, or refer to a higher-level*. (P7_HCW_FGD02_KLF)

While training enhanced CSWs’ ability to identify children and HCWs’ capacity to assess and manage DDs, both groups continued to experience role-specific gaps, CSWs in responding to detailed caregiver questions, and HCWs in confidently managing complex cases and ensuring diagnostic accuracy.

A CSW highlighted:

> *I went to the household, and a caregiver asked me what kind of food a child with autism should eat. So, when you are asked such questions…we [CSW] did not know what to say*. (P4_CSW_FGD01_NBO)

A HCW recounted uncertainty in managing behavioural or high-risk cases:

> *The child became violent… and I felt like I did not help as much as I should have because I did not know what to do…I was asking myself…do we restrain the child? What do we do next?* (P9_HCW_FGD02_KLF)

Another HCW reflected on challenges in diagnostic decision-making:

> *We had about three children come in, and we diagnosed them with autism, but later it turned out to be a misdiagnosis*. (P11_HCW_FGD05_KLF)

Participants from both cadres reported that the training, while valuable, was insufficient in duration and depth to fully prepare them for real-world practice. HCWs described the training as intensive but too short to cover the complexity of DDs:

> *Okay, I would like to say the training was good. But… if you were to train again, increase the number of days because it was a lot of information that was being disseminated…if somebody has never been trained on this, they would not grasp anything*. (P6_HCW_ FGD03_NBO)
>
> *The 3 days were quite packed and intense; at least it should have taken like 5 days*. (P11_HCW_FGD04_NBO)

### Caregiver engagement, community awareness, myths, and misconceptions on DDs

Beyond individual competencies, participants reported improved capacity to engage caregivers and communities, including addressing myths, misconceptions, and negative beliefs regarding DDs and promoting early help-seeking.

#### Caregiver education and counselling

Both CSWs and HCWs described that the training enhanced their capacity to educate caregivers about DDs, emphasising the importance of early intervention and dispelling misconceptions.

> *We advise parents that early support helps the child improve, and we explain that the child can still learn and grow*. (P6_CSW_FGD05_KLF)

HCWs noted that many caregivers lacked understanding of their children’s conditions, with some attributing symptoms to supernatural causes. They also educated the caregivers about the DD diagnosis and counselled them.

> *Some parents didn’t know that the children had these disorders… some thought maybe it was witchcraft. We educated the parents about what it is and referred them to places where they could get help*. (P1_HCW_FGD01_NBO)

Participants also shared that the training created broader community-level awareness that extended beyond individual consultations.

> *Everyone [in the community] is now curious, if my child delays, I know I have to do something*. (P4_HCW_FGD01_NBO)

#### Dispelling myths and misconceptions on DDs

Both cadres described how their ability to address misconceptions about DDs during routine interactions with caregivers and communities had improved. A CSW highlighted how the training enabled them to challenge these beliefs:

> *Before, people believed these children were affected by witchcraft, but now we can explain clearly and reduce that fear*. (P7_CSW_FGD04_NBO)

HCWs described how increased knowledge allowed them to reframe caregiver understandings and provide alternative explanations grounded in health:

> *Some parents used to hide their children, and they used to believe this was witchcraft, but by educating them about health and addressing them*. (P7_HCW_FGD02_KLF)

At an individual-level, HCWs also reflected on changes in their own beliefs:

> *It gave me a different mind towards those people who have disability… they are one of us*. (P1_HCW_FGD05_NBO)

#### Respectful, informed communication and engagement

Both cadres shared that they gained skills to respectfully and empathetically engage with families. Participants emphasised the importance of appropriate language and non-judgemental approaches.

> *We were taught not to call children ‘disabled’ but to say children with disability, because the words we use can make parents feel hopeless*. (P6_CSW_FGD05_KLF)

HCWs similarly emphasised the need for patient-centred, compassionate communication.

> *You cannot see these people within a minute… you need to have a cool time with them, talk with them nicely, then explain to them what is happening*. (P2_HCW_FGD01_NBO)

#### Knowledge sharing with peers, Continuous medical education (CME), and the community

Both cadres acknowledged the impact of cascading the knowledge and skills gained from the training to other colleagues and community members. Some CSWs reported on how other peers and caregivers supported them in identifying children’s DDs, creating ripple effects of awareness.

> *When you educate one mother, she tells another, and slowly the community begins to understand [about DDs]*. (P7_CSW_FGD05_KLF)

HCWs also highlighted their acquired capacity to sensitise other HCWs in the facility to continue identifying children with DDs.

> *We shared what we learnt during CMEs, and gradually, other staff became more aware of how to identify children who need further assessments*. (P8_HCW_FGD05_KLF)

Although training enabled participants to challenge misconceptions and improve awareness, entrenched beliefs and initial resistance shaped community engagement in some contexts. CSWs highlighted that engagement with caregivers was often difficult at first, requiring repeated visits before trust could be established:

> *Some caregivers in the community hold stigmatising attitudes, so when you approach them for the first time, there is often resistance, and they may not receive you. However, with continued effort, by the second or third visit, you begin to build rapport*. (P12_ CSW_FGD03_NBO)

Persistent misconceptions were further reflected in caregiver denial of developmental conditions during interactions. Acceptance of children’s conditions was not always immediate, requiring ongoing explanation and engagement.:

> *There are caregivers living with these children who insist that the child has no problem at all*. (P1_CSW_FGD04_NBO)

Similar challenges were observed at the facility-level, with caregivers continuing to attribute DDs to non-medical causes, with concealment of children still hindering effective identification and linkage to care:

> *These children [with DDs] were being hidden… the parent hides these children*. (HCW, FGD03_NBO)

### Strengthening referral pathways, system linkages and continuity of care

Participants described how training-related gains extended to broader service delivery processes, including referral systems, intersectoral collaboration, and continuity of care. However, these improvements were accompanied by implementation challenges.

#### Improved referral decision-making

CSWs shared how the training provided a better understanding of the locally available referral process, while HCWs used the knowledge gained to improve protocols for specialised services.

> *We understood the difference between delayed milestones and disability, and for those who have not been assessed, we refer them to the facility*. (P12_CSW_FGD01_NBO)
>
> *We became more aware of where to send children depending on their needs. Before, referrals were not very clear, but now we know which centres can help with assessment or therapy*. (P4_HCW_FGD04_NBO)

#### Strengthening community health facility linkages

Both CSWs and HCWs described the role of the training in strengthening the link between community and facility levels. Some HCWs recognised the importance of CSWs in early identification and referral.

> *The CHPs [a cadre of the CSWs] played a big role because, once they understood what to look out for, they referred children to the facility earlier*. (P6_HCW_FGD04_NBO)

Other CSWs shared that the referral pathways felt more functional and effective after the training.

> *The linkage with the health facility made referrals more than just paperwork; there was someone ready to receive the child.* (P4_CSW_FGD05_KLF)

#### Multidisciplinary teamwork and peer collaboration

Both cadres reported improved team-based approaches across multiple disciplines, such as rehabilitation, nutrition, maternal and child health (MCH), and education, post-training.

> *We worked as a team, went back to our notes, discussed, and came up with a conclusive diagnosis*. (P6_HCW_FGD03_NBO)
>
> *Even though not everyone was trained, we worked as a team. When a child came in, we involved colleagues from nutrition and MCH to support the assessment and follow-up*. (P2_HCW_FGD04_NBO)
>
> *Teamwork helped; we identified children from school, home, and clinic*. (P2_CSW_FGD02_KLF)

Participants described a collaborative working environment that supported client care and strengthened service delivery:

> *There was very good teamwork… seeing my colleagues work with those clients*. (P10_HCW_FGD02_KLF)

They also highlighted the practical value of peer assistance in managing workload and stress during service provision.

> *When I was overwhelmed, somebody from the team came for assistance, so I felt supported*. (P11_HCW_FGD02_KLF)

#### Continuity of care and follow-up

Both cadres expressed taking more proactive efforts to trace families of children with DDs who defaulted from care, reminding them of the importance of sustained engagement.

> *When a caregiver misses follow-up, we visit their home to remind them why clinic visits are important*. (P9_CSW_FGD01_NBO)

#### Implementation challenges

Despite strengthened referral linkages and improved collaboration across cadres, participants described several constraints that shaped implementation. An increased workload, in addition to the already high routine workload prior to the study, was reported by participants.

> *As the number of referrals increased, managing them became difficult because we had only a few staff members, and developmental assessments took time*. (P5_HCW_FGD04_NBO)

This pressure was made worse by the already high number of patients seen in health facilities:

> *Most of the time we see about 130 to 150 patients per day, but since the research started, the number has increased because we are also receiving more children with developmental issues*. (P3_HCW_FGD01_NBO)

CSWs described competing demands between routine duties and follow-up responsibilities.

> *The work became heavy because we were expected to do our usual community duties and still add child follow-up and referrals*. (P7_CSW_FGD03_NBO)

Additional challenges included limited availability of specialists, which limited effective referrals for further support, weak referral feedback mechanisms, where specialists did not communicate assessment outcomes to HCWs, and transport costs for families, also constrained referral uptake:

> *You advise them to go to the hospital, but they often lack transport money, so you have to support them financially or find ways to help them access the service*. (P2_CSW_FGD03_NBO)

Both CSWs and HCWs said that one-off training was insufficient and that ongoing learning and supervision were critical to support implementation. The small number of trained participants was also reported to have limited coverage, and expanded capacity building across cadres was recommended:

> *In XXX we have 200 CHPs [a cadre of CSWs] … only about 30 participated… maybe more CHPs should be trained so they can cover more households*. (P6_CSW_FGD04_NBO)
>
> *I would recommend we train more health care workers since not everybody has this knowledge*. (P5_HCW_ FGD04_NBO)

Participants also described the emotional and relational complexity of their roles, particularly when working with vulnerable households:

> *You go into a household, and the caregiver tells you they have not eaten for days… so before you even begin the assessment, you have to first address that situation*. (P3_CSW_FGD03_NBO)

These situations required empathy, trust-building, and flexibility, often extending beyond formal role expectations.

Across themes, strengthened roles in the identification, referral, assessment, and management of children with developmental delays were supported by training and intersectoral collaboration. Important to note is that while there were important shifts, health system constraints and barriers impacted the implementation experiences and the extent to which CSWs and HCWs could implement the lessons learnt.

## Discussion

This mixed-methods study assessed a capacity-strengthening intervention for community and facility-based frontline providers designed to enhance early identification, referral, and support for children with DDs in Kenya. The findings demonstrate that the training format was aligned with their needs, preferences, and norms/expectations. The training improved DD-related knowledge among HCWs and increased confidence, role clarity, and perceived confidence across both cadres.

Consistent with evidence from other LMICs, our findings suggest that gaps in early identification were primarily related to limited frontline capacity, weak referral systems, and inadequate supervision [1, 40–42]. Most participants reported no prior training in DDs, and, when present, previous exposure focused narrowly on assessment rather than on caregiver engagement or linking families to services. Similar gaps in developmental and child mental health training among frontline providers have been widely documented across LMICs, where non-specialist health workers often encounter developmental concerns but receive little structured preparation to identify or manage them [23, 43]. Uncertainty in differentiating conditions with overlapping presentations, such as autism, ADHD, and learning or behavioural difficulties, was common, reflecting challenges widely reported in global DD and child mental health literature [20, 44, 45]. In this context, structured capacity building initiatives, such as those based on the WHO mhGAP, have increasingly been used to equip non-specialist providers with practical guidance for identifying and managing developmental and mental health conditions within primary care systems [43, 46].

Post-training, HCWs demonstrated statistically significant improvements in DD-related knowledge scores. Although absolute gains were modest, this pattern is consistent with short, foundational training interventions and reflects the complexity of DD knowledge acquisition [26–28]. Evidence from evaluations of mhGAP-based and other non-specialist training programmes similarly shows that short courses often produce relatively modest improvements in test-based knowledge, particularly when addressing complex neurodevelopmental conditions unfamiliar to general health workers [43, 46]. Variation in baseline knowledge, particularly among participants with prior exposure to related training, may also limit the magnitude of measurable change due to ceiling effects [47]. Importantly, qualitative findings indicate that these gains translated into greater clinical confidence and a willingness to act, suggesting that knowledge served as an enabling mechanism rather than an endpoint. This aligns with the task-sharing literature, which shows that the early impacts of training are often strongest on self-efficacy and role legitimacy [21, 22, 48], though this training alone is not enough and can instead form part of planned, more intense training to enhance care.

A key contribution of this study is evidence that training both community and facility-based cadres strengthens the identification-to-referral process. CSWs acted as early identifiers and navigators, supporting caregiver engagement, referral uptake, and sustained treatment, while HCWs provided diagnostic oversight and clinical management. This dual-tier approach reduced division between community and facility services, addressing a common limitation of single-cadre training models in LMICs [8, 49, 50]. Evaluations of mhGAP implementation and other task-sharing programmes have similarly highlighted the importance of integrating community-level actors with facility-based providers to improve case detection, referral continuity, and service uptake [46, 51].

CSWs also reported a shift from focusing on impairments with visible physical symptoms to recognising neurodevelopmental and behavioural conditions, challenging entrenched assumptions about disability. Similar changes in conceptualisation have been observed in community-based DD interventions and are associated with earlier help-seeking and reduced stigma [13, 52–54]. Equipping CSWs with respectful, culturally appropriate language further supported engagement with caregivers, a critical determinant of early identification. Caregiver studies across LMICs and other settings, which document prolonged delays between initial recognition of concern and diagnosis [16]. The findings underscore that early identification is not solely a technical task but a relational process requiring communication, trust, and navigation support. Training that strengthened caregiver-facing skills was perceived to improve referral follow-through, aligning with evidence supporting family-centred approaches to developmental care [30, 55, 56].

Participants reported cascade effects, with trained CSWs and HCWs sharing knowledge through peer mentorship, CHPs, and continuing medical education sessions. Such diffusion has been reported in task-sharing and cascade training models and is increasingly recognised as important for scalability in resource-constrained settings [8, 49, 57]. Although downstream effects were not measured, these findings suggest that embedding training within existing community and health system networks may extend reach beyond direct trainees.

Despite these gains, implementation was constrained by staff shortages, high workloads, limited availability of specialists, transport costs for families, and weak referral feedback mechanisms. These barriers reflect well-documented health system constraints in LMICs [58, 59]. Importantly, these challenges reflect health system limitations rather than lack of motivation among frontline providers, consistent with evidence from DDs research showing that under-identification and poor follow-up in LMICs are driven primarily by weak system integration, limited supervision, and fragmented referral pathways rather than provider unwillingness [21, 60, 61].

### Policy and practice implications

These findings support a shift from facility-centric, single-cadre training models toward integrated, multi-cadre approaches embedded within community and PHC systems. Crucially, this requires a multisectoral orientation that formally incorporates teachers, community health workers, and caregivers alongside PHC providers within early identification and care pathways. Priorities include routine community-and school-linked screening supported by ongoing supervision, clear task-sharing, referral, and accountability across cadres and sectors; and meaningful caregiver engagement in identification, care decisions, and ongoing management. Equally important is aligning capacity-strengthening efforts with functional referral systems, including bidirectional communication and feedback loops among community, education, and health services. Without such coordination, early identification gains are unlikely to translate into timely access to care. These priorities align with global calls to strengthen frontline systems to meet early childhood development targets and reduce inequities in access to developmental services [30].

### Strengths and limitations

Strengths include the mixed-methods design, inclusion of multiple cadres, and triangulation of quantitative and qualitative findings. Limitations include reliance on self-reported data and a geographic focus on two counties, which may limit generalizability. Nonetheless, the convergence of findings strengthens their interpretation and transferability to other similar contexts. Future research should examine the longitudinal effects of training through repeated exposure models, including refresher sessions, structured supervision, and peer learning, to assess their role in sustaining knowledge retention and progressively strengthening provider competence in complex domains such as developmental and child mental health care [43, 48, 62].

## Conclusion

This study shows that multi-tiered capacity-strengthening of CSWs and primary HCWs can improve knowledge, confidence, and early identification practices for children with DDs in LMICs. Training strengthened community-facility linkages and dispelled myths and misconceptions on DDs but was insufficient to overcome entrenched system barriers. Sustained gains will require ongoing supervision, stronger referral systems, and cross-sectoral collaboration. Community-facility partnerships offer a feasible pathway to reducing diagnostic delays and advancing equitable, family-centred developmental care.

## Author contributions

AA, RAH, and CN contributed to the conception, design, and funding acquisition, with funding awarded to AA and RAH as SPARK co-principal investigators. AA, RAH, CN, VA, MWN, MD, THK, FG, BM and EM contributed to the protocol development and methodology. The development of training content was supported by members of the SPARK consortium from Ethiopia, Kenya, and the UK, including BM, VA, EM, THK, FG, MD, MWN, and RAH. BM, VA, EM, CN, and NK provided various support during the mhGAP and CSW training in Kilifi and Nairobi. WK, EC, and BM provided clinical input during HCW trainings and post-training implementation, with technical support from FGM. Data collection and curation were supported by BM, EM, CN, and NK with supervision from VA. BM, VA, SN, EM, CN, AAA, CN, and MS were involved in various phases of data analysis and interpretation. BM wrote the first draft and received technical input from VA, AA, CN, MS, RAH, MWN, and EM in subsequent versions of the manuscript. All authors contributed to the manuscript revisions, read, and approved the submitted manuscript.

## Acknowledgements

We thank all study participants for their contribution to this research. We acknowledge the support from Kilifi and Nairobi (sub)county health management teams, as well as SPARK’s community and national project advisory board members. We thank all stakeholders and workshop participants in Kenya and Ethiopia for their contributions to the development of training content. We acknowledge SPARK team members for their support in training, community engagement, and data collection: Brenda Nzioka, Eunice Ombech, Victoria Lewa, Ann Mumira, Veronica Achoki, Constance Yaa, Joy Yvonne Thoya, Ann Muisyo, Damaris Nguyo, Kevin Sidika, Derrick Otieno, Patrick Idd, Charo Ngoka, Richard Charo, Khamisi Katana, and Beatrice Kabunda.

## Data availability statement

Data cannot be shared publicly because it contains potentially identifying or sensitive information about study participants. Data are available upon request to qualified researchers, subject to approval, from the ISERC at the Aga Khan University.

## Supporting information captions

S1 Table. Sociodemographic characteristics of the qualitative sample: Community Support Workers

S2 Table. Sociodemographic characteristics of the qualitative sample: Healthcare Workers S3 Table. Themes, sub-themes, from HCWs and CSWs perspectives on DD training

